# Differentiating the Effect of Medication and Illness on Brain Volume Reductions in First-Episode Psychosis: A Longitudinal, Randomized, Triple-blind, Placebo-controlled MRI Study

**DOI:** 10.1101/2020.03.18.20038471

**Authors:** Sidhant Chopra, Alex Fornito, Shona M. Francey, Brian O’Donoghue, Vanessa Cropley, Barnaby Nelson, Jessica Graham, Lara Baldwin, Steven Tahtalian, Hok Pan Yuen, Kelly Allott, Mario Alvarez-Jimenez, Susy Harrigan, Kristina Sabaroedin, Christos Pantelis, Stephen J Wood, Patrick McGorry

**Affiliations:** Monash University, Turner Institute for Brain and Mental Health, School of Psychological Sciences and Monash Biomedical Imaging

**Author notes:** **Authors contributed equally** (Sidhant Chopra, Alex Fornito). **Corresponding Author:** Alex Fornito, Turner Institute of Brain and Mental Health, 770 Blackburn Road, Clayton, Victoria, Australia.

## Abstract

Changes in brain volume are a common finding in Magnetic Resonance Imaging (MRI) studies of people with psychosis and numerous longitudinal studies suggest that volume deficits progress with illness duration. However, a major unresolved question concerns whether these changes are driven by the underlying illness or represent iatrogenic effects of antipsychotic medication. Here, we report MRI findings from a triple-blind randomised placebo-controlled study where 62 antipsychotic-naïve patients with first episode psychosis (FEP) received either an atypical antipsychotic or a placebo pill over a treatment period of 6 months. Both FEP groups received intensive psychosocial therapy. A healthy control group (n=27) was also recruited. Structural MRI scans were obtained at baseline, 3-months and 12- months. Our primary aim was to differentiate illness-related brain volume changes from medication-related changes within the first 3 months of treatment. We secondarily investigated long-term effects at the 12-month timepoint. From baseline to 3 months, we observed a significant group x time interaction in the pallidum (p < 0.05 FWE-corrected), such that patients receiving antipsychotic medication showed increased volume, patients on placebo showed decreased volume, and healthy controls showed no change. In patients, a greater increase in pallidal grey matter volume over 3 months was associated with a greater reduction in symptom severity. We additionally found preliminary evidence for illness- related volume reductions in prefrontal cortices at 12 months and medication-related volume reductions in cerebellum at both 3-months and 12-months. Our findings indicate that psychotic illness and antipsychotic exposure exert distinct and spatially distributed effects on brain volume. Our results align with prior work in suggesting that the therapeutic efficacy of antipsychotic medications may be primarily mediated through their effects on the basal ganglia.

## Introduction

Magnetic Resonance Imaging (MRI) has been used extensively to document brain changes in psychotic disorders. Grey matter volume (GMV) reductions relative to healthy controls are particularly robust, and evident across all illness stages^1 2 3^ and in multiple brain regions^4 5 2^. Some of these changes appear to worsen with transition to psychosis and ongoing illness^6^, which has been taken as evidence of a progressive process associated with illness onset^7^, although some have opposed this view^8,9^.

Numerous mechanisms have been proposed to explain longitudinal brain changes in schizophrenia, including aberrant neurodevelopment^10^, neuroinflammation^11^, network-based pathological spread^12^, and the iatrogenic effects of antipsychotic treatment^4,13^. In particular, widespread and early treatment of patients with antipsychotics has made it notoriously difficult to disentangle the effects of medication and pathophysiology on brain volume. Although studies of antipsychotic-naïve patients clearly show brain GMV reductions in the absence of medication^2^, several lines of evidence suggest that antipsychotic medication influences GMV^14^. For example, longitudinal studies suggest that cumulative exposure to antipsychotic medication is associated with reduced total cerebral^13^ and prefrontal GMV^4^, and studies in macaques have shown that chronic exposure to typical and atypical antipsychotics reduces total GMV^15^ and glial cell number^16^. One recent placebo-controlled trial in mostly remitted patients with psychotic depression showed that, compared to patients on placebo, those given olanzapine had decreased cortical thickness within both hemispheres^17^.

Other work suggests that antipsychotic medication, particularly atypicals, may exert a neuroprotective effect^18^. Studies in rodents have supported a neuroprotective effect of atypicals^19^, which may arise through several candidate mechanisms, including neurogenesis ^19^ and protection against oxidative stress^21^. This work parallels naturalistic^22^ and experimental^18^ longitudinal MRI studies in human patients suggesting that atypical antipsychotics may be associated with less GMV loss when compared to typical antipsychotics.

One limitation affecting all existing longitudinal studies conducted thus far is that they have only examined patients who are receiving antipsychotic medication. This approach is problematic because previous exposure to medication may result in brain changes that could mask or be mistaken for illness-related processes. The only way to unambiguously distinguish illness-related from medication-related brain changes is through a randomized placebo-controlled study of antipsychotic naïve first-onset patients, in which one patient group is exposed to antipsychotic medication and the other receives a placebo. This design is able to test several distinct hypotheses about the differential contributions of illness and antipsychotics to brain changes in the earliest illness stages (Figure 1). However, such experiments are difficult to conduct due to the practical difficulties and ethical concerns associated with withholding antipsychotic treatment.

**Figure 1.**
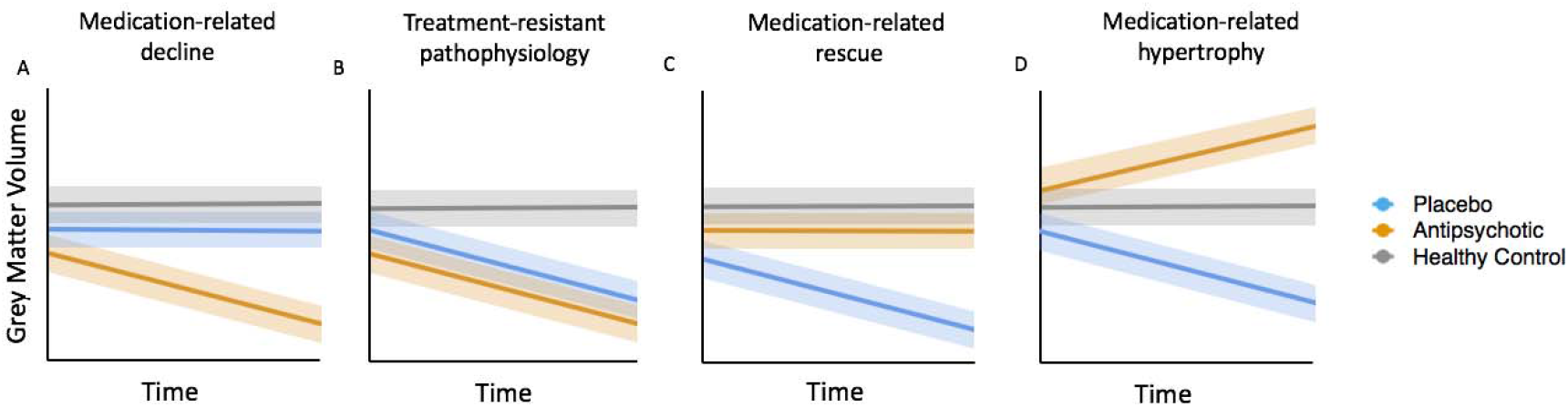
*Disentangling illness-related and antipsychotic medication-induced brain changes in early psychosis using a randomized placebo-controlled design*.

We recently overcame these challenges to conduct, to our knowledge, the first randomized, triple-blind, placebo-controlled trial of antipsychotic medication in first-episode psychosis (FEP), in which antipsychotic-naïve patients were randomized to receive psychosocial therapy with or without antipsychotic medication over the first six months of treatment engagement^23^. We found, using a non-inferiority design^23^, that the placebo group showed comparable clinical and functional outcomes to the medicated group at the end of the treatment study^24^. Here, we report an analysis of GMV in this cohort, where MRI was acquired before treatment (baseline), at three months, and then at a 12-month follow-up. Our primary aim was to distinguish volumetric changes attributable to illness from those attributable to antipsychotic medication within the initial 3-month period (Figure 1). Our secondary aim was to investigate longer-term changes at the 12-month follow-up, after a period of time in which both groups had been exposed to antipsychotics. We also examined whether any observed volumetric changes were associated with symptomatic and functional changes.

Each panel presents a schematic of expected results under different hypotheses. (A) A medication-related decline due to antipsychotics is indicated if medicated patients show accelerated GMV loss compared to patients in the placebo group and healthy controls. (B) An illness effect that is not modified by treatment is indicated if both treatment groups show accelerated GMV loss relative to controls. (C) An illness-related change that is rescued by antipsychotics is indicated if GMV loss is observed in the placebo group but not medicated patients. (D) Antipsychotic-related hypertrophy, where GMV is increased in the medicated group compared to the healthy controls and/or placebo group, could be consistent with either a possible medication-related rescue or the initial stages of a volume-loss process (e.g., an oedemic reaction). These possibilities could be disentangled by examining correlations with symptomatic or functional measures; e.g., an association between the volumetric increase and improved outcome would be consistent with possible rescue. For simplicity, controls are depicted as showing no change over time, but they may also show longitudinal increases or decreases. The key factor is whether the rate of change is greater in patients compared to controls. Solid lines represent group means and shaded areas represent some estimate of the error around the mean.

## Method

### Study Design

Patients were randomized to one of two groups: one given antipsychotic medication plus intensive psychosocial therapy (MIPT) and the other given a placebo plus intensive psychosocial therapy (PIPT) (Figure 2). A third healthy control group who received no intervention was also recruited. For both patient groups, the treatment period spanned six months. MRI and clinical assessments were conducted at baseline, three months, and a final follow-up at 12 months. The randomization phase of the study terminated at 6 months, so patients in either the MIPT or PIPT group could have received antipsychotic medication and ongoing psychosocial interventions in between the 6 and 12 months into the study. Further research and safety protocols can be found in the Supplement and elsewhere^23^.

**Figure 2.**
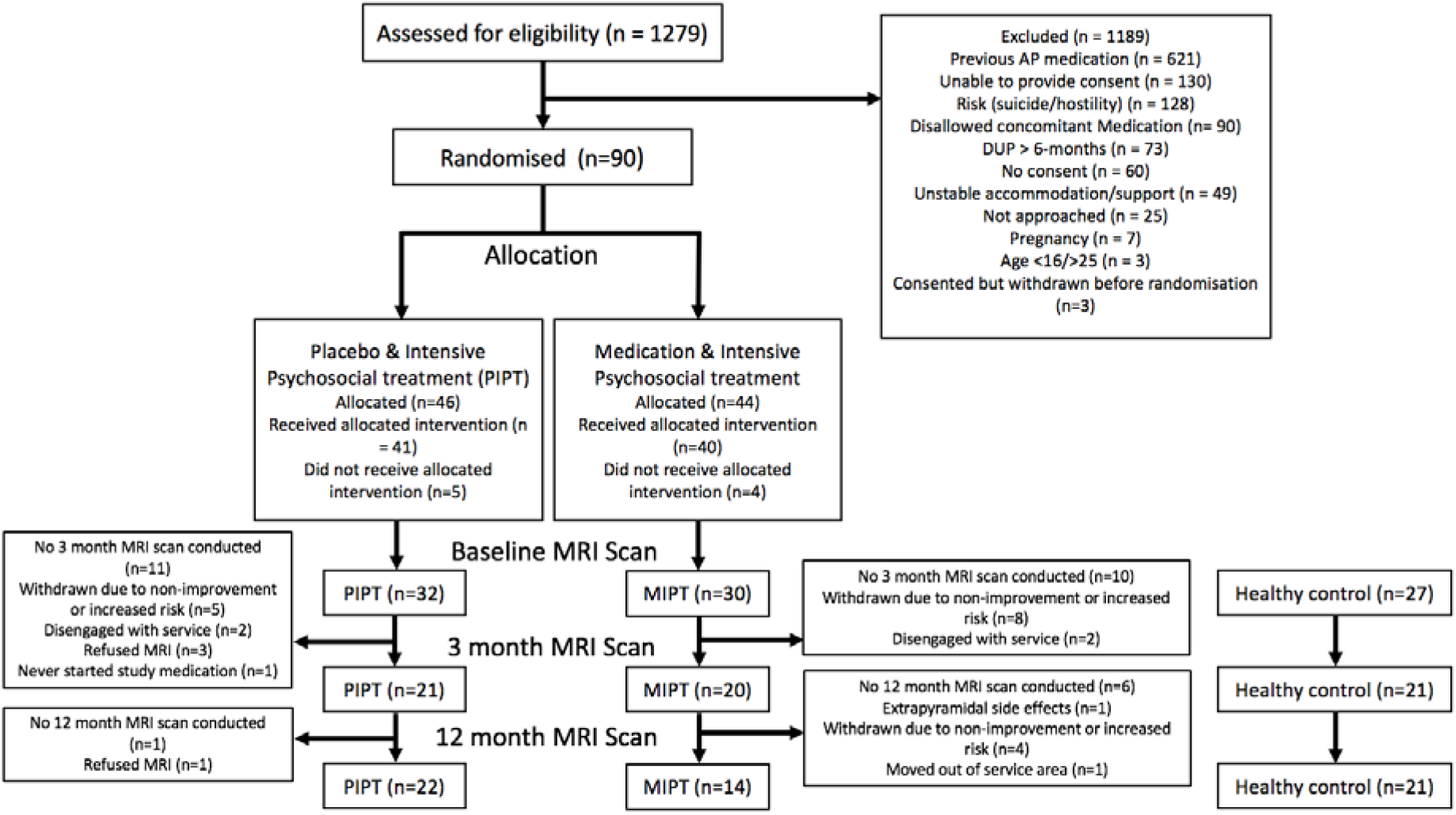
– Recruitment flow diagram for the patient group.

### Participants

Patients were aged 15-25 yrs and were experiencing a first episode of psychosis, defined as fulfilling Structured Clinical Interview for DSM-5 (SCID) criteria for a psychotic disorder, including schizophrenia, schizophreniform disorder, delusional disorder, brief psychotic disorder, major depressive disorder with psychotic symptoms, substance-induced psychotic disorder or psychosis not otherwise specified. Additional inclusion criteria to minimise risk were: ability to provide informed consent; comprehension of English language; no contraindication to MRI scanning; duration of untreated psychosis (DUP) of less than 6 months; living in stable accommodation; low risk to self or others; minimal previous exposure to antipsychotic medication (< 7 days of use or lifetime 1750mg chlorpromazine equivalent exposure; further details provided in Supplementary Table 1).

Healthy control participants were aged between 18 and 25, could provide written informed consent, and were psychiatrically, neurologically and medically healthy. A stratified randomisation design, with Gender and DUP as factors, was used to allocate patients to either MIPT or PIPT treatment groups. DUP was included as a three-level factor (0-30 days, 31-90 days, and >90 days). Clinicians, patients, study assessors and researchers conducting MRI pre-processing remained blinded to treatment allocation throughout the trial. Further details on inclusion criteria, safety measures, and discontinuation criteria can be found elsewhere^23^. A recruitment flow diagram and final group numbers at each time point are presented in Figure 2 and demographic are presented in Table 1.

**Table 1.** – Sample characteristics and group differences at baseline. Abbreviations: 245 PIPT = placebo plus intensive psychosocial therapy, MIPT = antipsychotic medication plus intensive psychosocial therapy; NOS = not otherwise specified; BPRS = Brief Psychiatric Rating Scale version 4; SOFAS = Social and Occupational Functioning Assessment Scale. SANS = Scale for the Assessment of Negative Symptoms; HAM-D = Hamilton Depression Rating Scale; HAM-A = Hamilton Anxiety Rating SCALE; QLS = Quality of Life Scale. ^†^ This column provides the T or χ^2^ values comparing the healthy control and patients (collapsed across two treatment conditions) at baseline.

| | First episode psychosis | | Healthy control (N=27) | T / $\chi^2$ (p) <sup>†</sup> |
| --- | --- | --- | --- | --- |
|  | PIPT<br>(N =30) | MIPT<br>(N=29) |  |  |
| Baseline age, years (SD) | 18.8 (2.72) | 19.5 (2.94) | 21.9 (1.93) | -4.49 (<0.001) |
| Females, N (%) | 14 (46.6%) | 13 (44.8%) | 17 (62.9%) | 2.19 (0.139) |
| Handedness, Left , N (%) | 2 (6.7%) | 2 (6.9%) | 3 (11.1%) | 0.465 (0.495) |
| Education, years (SD) | 11.8 (1.86) | 12.7 (2.30) | 15.2 (1.90) | -6.21 (0.001) |
| Diagnosis, N |  |  |  |  |
| Major depression with psychosis | 7 | 5 | - |  |
| Schizophreniform disorder | 5 | 5 | - |  |
| Psychotic disorder NOS | 8 | 7 | - |  |
| Substance-induced psychotic disorder | 4 | 2 | - |  |
| Delusional disorder | 1 | 4 | - |  |
| Schizophrenia | 5 | 5 | - |  |
| Missing diagnosis | 0 | 1 | - |  |
| Baseline BPRS Total, mean (SD) | 59.4 (9.64) | 55.8 (10.10) | - | 1.40 (0.166) |
| Baseline SOFAS, mean (SD) | 52.9 (14.0) | 51.7 (10.6) | - | 0.384 (0.703) |
| Baseline SANS, mean (SD) | 37.3 (17.2) | 32.9 (18.1) | - | 0.961 (0.346) |
| Baseline HAM-D, mean (SD) | 19.5 (6.75) | 18.2 (6.57) | - | 0.745 (0.459) |
| Baseline HAM-A, mean (SD) | 22.2 (6.42) | 20.2 (7.25) | - | 1.118 (0.268) |
| Baseline QLS, mean (SD) | 68.5 (24) | 70.6 (20.8) | - | -0.368 (0.714) |

### Symptomatic and functional measures

The preregistered primary and secondary trial outcome measures were the total Social and Occupational Functioning Assessment Scale (SOFAS) and the BPRS-4 scores respectively^23^. Additional measures can be found in Supplementary materials.

### Antipsychotic Medication

Patients randomised to the MIPT group received either 1mg risperidone (n=25) or 3mg paliperidone (n=5). To reflect real-world clinical treatment, this starting dose was then increased according to clinical response by the blinded treating clinician. The same procedure was followed for participants in the PIPT group, who received a placebo pill that was identical in taste, appearance, and packaging to the active medication. Additional details can be found in Supplementary materials.

### MRI acquisition and pre-processing

A 3-T Siemens Trio Tim scanner located at the Royal Children’s Hospital in Melbourne, Australia, was used to acquire a high resolution structural T1-weighted scan for each participant. Pre-processing of scan was done using the Computational Anatomy Toolbox and Diffeomorphic Anatomical Registration Exponentiated Lie algebra algorithm (DARTEL)^25^. Additional details can be found in Supplementary materials.

### Statistical analyses

Mixed effects marginal models were used to analyse regional GMV across the three groups (MIPT, PIPT and Healthy control) and three-time points (baseline, three-months and 12- months follow-up). The models were implemented at voxel-level in the Sandwich Estimator Toolbox^26^ (version 2.1.0). All other statistical analyses were conducted in R-studio (version 1.1.423).

Our primary analyses sought to disentangle the effects of medication and illness (e.g., Figure 1) on total GMV and to map localised changes using VBM. This analysis focused on the baseline and 3-month timepoints, as they fell within the treatment period. For both total and regional GMV, we first tested for baseline differences between groups using an analysis of covariance (ANCOVA), controlling for age at baseline, sex, handedness and total intracranial volume. We then examined longitudinal changes using a marginal model. The voxel-level analysis was implemented in the Sandwich Estimator Toolbox^26^ (version 2.1.0), which uses ordinary least squares estimators of group-level regression parameters and a modified sandwich estimator for standard errors^27^. This method allows for robust and accurate estimation of random effects while mitigating problems posed by mis-specification of covariance structure when using traditional mixed-effects models^26^. The contrast of interest was a group (MIPT, PIPT, Control) by time (baseline, 3-month) interaction. We performed inference using non-parametric bootstrapping (10,000 bootstraps), with statistical significance for total volume assessed at p <0.05, and a p<0.05, family-wise error (FWE)- corrected threshold for voxel-level analyses, as implemented in the Sandwich Estimator Toolbox^26^. To provide a more complete picture, we also report results surviving a less stringent threshold of p<0.001, uncorrected, with an extent threshold of 10 voxels, but caution that these findings require replication. Our secondary analysis included the 12-month timepoint and was designed to examine the long-term effects of early withholding of antipsychotic medication. Similar procedures were used as in the primary analysis. Details of the 12-month analysis and of analyses addressing symptom correlates and confounding variables, are in the Supplement.

## Results

### Demographics and clinical characteristics

There were no significant differences between the patient and control samples in sex or handedness, but the patients were, on average, 1.9 years younger and had 2 years less education (Table 1). At baseline, the two patient groups (PIPT and MIPT) did not significantly differ in age, education, sex, handedness, BPRS or SOFAS score.

### Baseline differences in total and regional grey matter volume

No significant baseline difference in total GMV was detected between patients (collapsed across treatment groups) and healthy control participants (F = 0.297; p = 0.588), nor were any voxel-level regional differences detected following whole-brain FWE-correction. Results at an uncorrected threshold can be found in Supplementary Table 3.

### Disentangling medication-related and illness-related brain changes in the first three months of treatment

No significant group by time interaction in total GMV was detected between the three groups (F = 0.387, p = 0.689). Using VBM to map regional changes, a significant group by time interaction was identified within the right pallidum (p < 0.05, FWE-corrected; Figure 3a). From baseline to 3 months, GMV in this region remained stable in controls, decreased in the PIPT group, and increased in the MIPT group (Figure 3b). The primary post-hoc contrasts revealed that, compared to baseline, pallidal GMV significantly decreased in PIPT patients (t = 2.34, p = 0.021), increased in MIPT patients (t = -2.20, p = 0.029), and did not change in controls (t =-0.142, p = 0.888). Secondary post-hoc contrasts conducted at the 3-month timepoint showed that the PIPT had significantly less pallidal GMV than the MIPT (t = -2.26, p = 0.012), with neither group differing from controls.

**Figure 3.**
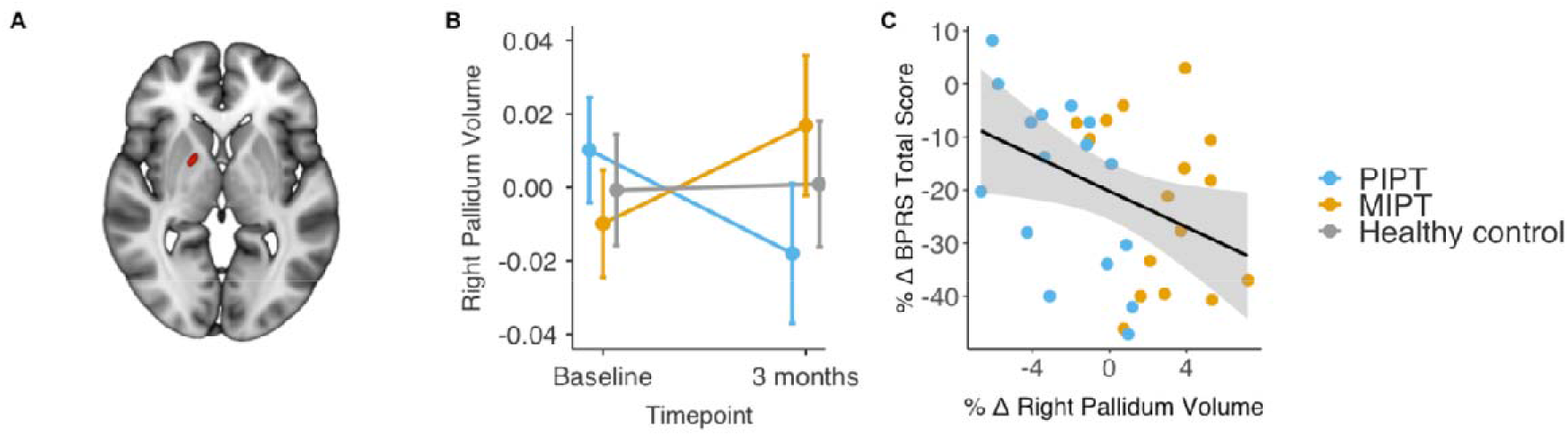
– **(A):** Red = Location of the cluster within the right pallidum where significant group x time interaction (p < 0.05, FWE-corrected) was detected. **(B):** The principal pallidal GMV eigenvariate for each group at baseline and 3-month follow-up, adjusted for model covariates. Error bars show 95% confidence intervals. (**C):** The association between percentage change (%Δ) in total Brief Psychiatric Rating Scale score (BPRS; y-axis) and percentage change in pallidal GMV volume within the two treatment groups.

Greater increase in pallidal GMV over 3 months was associated with a greater reduction in symptom severity, as indexed by the BPRS Total score (ρ = -0.418; p = 0.017; Figure 3c). There was no significant association with SOFAS total score (ρ = -0.002; p = 0.998), and none of the exploratory correlations with ancillary measures survived correction for multiple comparisons. At an uncorrected threshold, we found a negative correlation between right pallidal volume and BRPS positive symptom change scores that was comparable in magnitude to the association with BPRS total (ρ = -0.431; p= 0.012), suggesting that the relationship between pallidal volume and symptom change may be specifically related to positive symptoms.

At an uncorrected threshold (k > 10, p < 0.001), we identified interactions between group and time that were consistent with a unmodified illness-related effect within lateral occipital cortex (Supplementary Figure 2a, Figure 1b); a medication-related decline within cerebellum (Supplementary Figure 2b, Figure 1a); and medication-related hypertrophy within the inferior temporal cortex, precuneus, and orbitofrontal cortex (Supplementary Figure 2c-e, Figure 1d).

### Potential confounds

No statistically significant associations between percentage change in pallidal volume between baseline and 3 months and DUP, concomitant medication use, or substance use were detected. Details are in the Supplementary materials.

### Disentangling medication-related and illness-related brain changes in the first 12 months of treatment

We next considered MRI measures at the 12-month follow-up to differentiate the long-term effects of medication and illness. Patients retained at the 12-month follow up did not significantly differ in baseline age (t = 1.7858, p = 0.08), sex (χ^2^ = 0.087, p = 0.767), education (t = 0.652, p = 0.518), BPRS (t = -0.076, p = 0.940) or SOFAS (t = 0.780, p = 0.439) from those who did not complete the 12-month follow up scan.

No statistically significant differences in linear trend for total GMV over the 12-month follow-up period were detected (F = 1.60, p = 0.192). At the voxel-level, no statistically significant regional differences were identified at the FWE-corrected threshold. At p < 0.001 (k=10) uncorrected, there were differences in linear trend consistent with unmodified illness-related changes (Figure 1b) within the bilateral dorsolateral superior frontal gyrus (Supplementary Figure 3a-b), right superior orbito-frontal gyrus (Supplementary Figure 3c), middle orbito-frontal gyrus (Supplementary Figure 3d), and left superior medial frontal gyrus (Supplementary Figure 3e); a medication-related decline (Figure 1a) within the right cerebellar crus I (Supplementary Figure 3f); and medication-related hypertrophy (Figure 1d) within the right middle temporal gyrus (Supplementary Figure 3g), temporal pole (Supplementary Figure 3h) and cerebellar VIII (Supplementary Figure 3i). The results were largely consistent when eight individuals within the PIPT group who were exposed to antipsychotic medication between the 3- and 12-month scans were removed from the analysis (Supplementary Figure 1).

Additionally, we assessed whether the changes seen within pallidal cluster detected in the primary analysis persisted at 12-month follow-up. The differences between the three groups were not statistically significant (Supplementary Figure 4).

### Assessing the specificity of findings to grey matter

To assess the specificity of our findings to grey matter, we repeated the above primary and secondary analyses in white matter. For the primary analysis, we found a significant group by time interaction within a small area of the left cerebellar lobule IX white matter (k = 9, p < 0.05, FWE-corrected; Supplementary Figure 5a). From baseline to 3 months, white matter volume in this region increased in the controls (t = -2.34, p = 0.021), remained stable in the PIPT group (t = 0.216, p = 0.830), and decreased in the MIPT group (t = 2.239, p = 0.027). This pattern of results is consistent with medication-related volume loss (e.g., Figure 1a). The change in volume within this cluster was not correlated with change in BPRS-4 or SOFAS. Results at an uncorrected threshold can be found in Supplementary Materials.

## Discussion

We used a triple-blind, placebo-controlled randomized trial to disentangle the effects of medication and illness on GMV change within early stages of first episode psychosis. We found evidence of regionally heterogeneous effects associated with both illness and medication, with the most robust effect being an illness-related decline of pallidal GMV in the placebo group coupled with an antipsychotic-related increase in the medicated group. Consistent with a therapeutic benefit of the antipsychotic-induced increase in pallidal GMV, a greater volumetric change in this area was associated with a greater reduction in symptomology within the first three months of illness. Preliminary evidence (k=10, p < 0.001, uncorrected) for unmodified illness-related changes and medication-related decline were identified in visual cortex and cerebellum, respectively.

Our secondary analysis of long-term changes assessed at 12-month follow-up revealed preliminary evidence of an unmodified illness-related GMV reduction in prefrontal cortex, medication-related decline in cerebellum, and medication-related hypertrophy in temporal areas. Together, these results suggest that both psychotic illness and medication exposure exert distinct and spatially distributed effects on GMV, and converge with prior work in suggesting that the therapeutic efficacy of antipsychotic medications is primarily mediated through their effects on the basal ganglia^28^.

### Illness-related volumetric reductions in FEP

Pallidal volume in the PIPT group declined over the first three months of illness. This decline was not associated with substance use or concomitant medication, which would be consistent with an illness-related effect. In contrast, MIPT patients showed an increase in pallidal volume over time. Thus, antipsychotic medication appears to prevent or perhaps even reverse illness-related volume loss in this part of the brain.

Using less stringent criteria for significance, we found evidence for illness-related GMV reductions in visual cortex within the first three months of illness, and further reductions in prefrontal cortex by the 12-month time point. Frontal white matter reductions were also identified at the 12-month time point. These changes were observed in both the PIPT and MIPT groups, suggesting that they are unmodified effects of illness.

The pallidum is the primary output structure of the striatum, and disturbances of fronto-striato-thalamic circuits have long been implicated in the pathogenesis of psychosis^29^. The function of these circuits is heavily modulated by dopamine, and their disruption is apparent in diagnosed patients^30^, patients’ unaffected first-degree relatives^30^ and individuals experiencing an at-risk mental state for psychosis^31^. Functional connectivity within this circuit also correlates with the severity of psychotic-like experiences in non-clinical samples^32^. Thus, one hypothesis that may explain our findings is that altered signalling from the striatum triggers early volumetric loss in the pallidum^29,33^, which subsequently spreads to affect functionally-related prefrontal areas^34,35^. This interpretation aligns with evidence that smaller pallidal volume in anti-psychotic naïve patients is associated with more severe psychiatric symptomology^36^, and our own finding that increased pallidal volume over the first three months correlates with improved symptom outcome. Moreover, our finding of possible long-term reductions in prefrontal cortex grey and white matter may explain why prefrontal dysfunction is so commonly reported in patients with established illness. However, we caution that the precise mechanisms underlying volumetric changes in psychosis remain a topic of debate^8,9^. Here, we show that some of these changes cannot be attributed to medication and other confounding factors, but more subtle influences such as differences in hydration, physical and mental activity, and stress levels cannot be ruled out. Targeted mechanistic studies are required before we can draw strong inferences about pathophysiological mechanisms.

### Are antipsychotics neuroprotective?

The increase of pallidal volume seen in MIPT patients, together with the correlation between increased pallidal GMV and symptom improvement between baseline and three months, are consistent with a putative neuroprotective effect of atypical antipsychotic medication (Figure 1d). Larger pallidal volumes have been widely reported in medicated^37^ but not antipsychotic naive^38^ patients. Human studies have also shown reduced volume loss in patients taking atypical compared to patients receiving typical antipsychotic medication^18^, and work in animals indicates that atypicals can exert several neuroprotective effects^19^, including induction of neurogenesis^20^, and protection against oxidative stress^21^, as well as positive effects on cognition^39^. Our results are in line with this work and suggest that atypicals prevent illness-related volume loss occurring early in the illness. However, we caution that MRI is unable to identify a specific cellular mechanism that would support a neuroprotection hypothesis, and the molecular mechanisms by which atypical medications might protect grey matter structures in humans are poorly understood. Atypicals are characterized by relatively high affinities for both serotonin and dopamine receptors^40^. Patients in our study received risperidone or its molecularly similar active metabolite paliperidone. Both medications are antagonists for 5HT_2_ receptors in addition to showing high affinity for D_2_ receptors^41^. Rodent studies using risperidone have demonstrated cell proliferation^42^, increased levels of brain-derived neurotrophic factor^43^, and the promotion of antioxidant defence^44^. Thus, while our data suggest that antipsychotics may rescue or perhaps reverse illness-related decline of pallidal volume within the first three months of illness, and that this apparent preservation of volume is associated with improved symptom outcomes, further studies are required to elucidate underlying cellular and molecular mechanisms.

Notably, pallidal volume had normalized in both MIPT and PIPT patients by 12 months (Supplementary Figure 4). While it is possible that this normalisation reflects differences in illness characteristics between patients who did and did not complete the 12 months follow-up, we found no significant difference in baseline demographic and clinical characteristics between these two groups. Nonetheless, it is possible that patients completing the 12-month assessment followed distinct illness trajectories after enrolment into the study. An alternative explanation is that early pallidal changes reflect an acute illness effect with subsequent normalisation reflecting a compensatory process. It is also possible that intensive psychosocial therapy and engagement with clinical services was sufficient to normalize volumes^45^.

### Evidence of medication-related decline in grey matter volume

We found no evidence for antipsychotic-related decline in GMV at whole-brain-corrected thresholds. At the less stringent, uncorrected threshold, we observed consistent medication-related GMV reductions in the cerebellum at 3 months and at 12 months (Figure 1a). Similar results surviving whole-brain correction were observed in cerebellar white matter. While previous naturalistic^4^ and experimental^17^ studies have demonstrated an association between antipsychotic medication and loss of both total and hemispheric volume, our study is distinct in several important ways. First, our study included a placebo control and healthy control, allowing us to experimentally isolate the effect of atypical antipsychotic medication. Second, we examined patients during a relatively short time span of one year, while other studies have examined longer periods^4^. Third, the mean cumulative dosage of antipsychotic medication in our study, while still an effective dose^46^, is considered low. Fourth, all participants in our study were scanned with the same scanner, mitigating the potentially confounding effects of scanner differences. Finally, all patients within our study received an evidence-based psychosocial intervention which may have associated neuroprotective effects^45^. In light of these differences, our results could be interpreted as preliminary evidence of potential neurotoxicity in early illness stages that is predominantly expressed in the cerebellum. However, we note that there was no association between volume change within the cerebellum and change in functional or symptom outcome scores, in either grey or white matter. Longer-term follow-up would be required to determine the extent to which further volume-loss emerges with additional antipsychotic exposure.

### Strengths and limitations

The strengths of this study include a prospective randomised control trial design, antipsychotic naïve patients, triple blinding to treatment, and the inclusion of a healthy control group as a reference for characterizing normative change over time. We also used robust non-parametric inference to model longitudinal changes in GMV^26^. In order for this study to satisfy ethical concerns, our inclusion criteria meant that patients who entered the study posed low risk of harm to self or others, lived in stable accommodation, and had a short DUP. Additionally, patients who did not improve in clinical symptomology or functioning were removed from the trial, which contributed to attrition. It is therefore possible that our final patient cohort represents a sub-sample of individuals with a lower severity of illness, and that the changes we report here are a conservative estimate of those that would be observed in a more heterogeneous sample. However, we note that the mean baseline SOFAS scores of our patients were comparable to epidemiologically representative cohorts of FEP patients^47^ and that the mean baseline BPRS score of patients within our study (57.6) would classify them as ‘markedly ill’^48^. Additionally, there were no differences in baseline clinical or demographic characteristics between patients who did and did not complete the study.

Thus, prima facie, there are few obvious differences between our cohort and many FEP samples reported in the literature, beyond the strict safety requirements of our study. Nonetheless, we cannot rule out the possibility that patients who remained in the study have a form of psychotic illness that is perhaps less severe and/or progressive than those who did not complete. A final limitation is that we only examined risperidone and paliperidone. It remains to be seen whether our results generalize to other antipsychotic medications.

## Conclusion

Taken together, our results demonstrate that psychotic illness and antipsychotic exposure exert distinct and spatially distributed effects on brain volume, with the most robust effect being consistent with an antipsychotic-related rescue of pallidal volume changes in the early stages of treatment.

## Data Availability

Please contact the authors.

## Supplementary Materials

### Study Design and Funding – Additional Details

The trial took place at the Early Psychosis Prevention and Intervention Centre, which is part of Orygen Youth Health, Melbourne, Australia. The trial was registered with the Australian New Zealand Clinical Trials Registry in November 2007 (ACTRN12607000608460) and received ethics approval from the Melbourne Health Human Research and Ethics committee.

Role of Funding Sources: Janssen-Cilag partially supported the early years of this study with an unrestricted investigator-initiated grant and provided risperidone, paliperidone and matched placebo for the first 30 participants. The study was then funded by an Australian National Health and Medical Research Project grant # 95757. The funders had no role in study design, data collection, data analysis, data interpretation, or writing of this report. The corresponding author had full access to all of the data in the study and had final responsibility for the decision to submit for publication.

### Additional Clinical and Functional Measures

Additional measures of clinical and functional included the Scale for Assessment of Negative Symptoms (SANS), Hamilton Depression Scale (HAM-D) and Hamilton Anxiety Scale (HAM-A), and the World Health Organisation Quality of Life Scale -Brief (WHOQoL-BREF). Substance use was measured using the World Health Organisation Alcohol, Smoking and Substance Involvement Screening Test (WHO-ASSIST). Duration of untreated psychosis was measures using clinical interview.

### Antipsychotic and Concomitant Medication

Concomitant medications were permitted during the trial, except for additional antipsychotics or mood stabilisers (rates of concomitant medications use are provided in Supplementary Table 2). Four patients within the PIPT group were switched to open-label antipsychotic medication before the 3-month MRI scan and were excluded from the primary analysis. After termination of the randomization phase at 6 months, five patients in the PIPT group were exposed to antipsychotic medication between the 3-month and 12-month scan. We examined the impact of this exposure in our analysis of the 12-month data, as detailed below. Mean cumulative dose and rates of exposure for both patient groups at each timepoint are provided in Supplementary Table 1. Duration of untreated psychosis (DUP) was assessed using a clinical interview.

### MRI Acquisition and Pre-processing

A 3-T Siemens Trio Tim scanner located at the Royal Children’s Hospital in Melbourne, Australia, was used to acquire a high resolution structural T1-weighted Magnetisation-Prepared Rapid Gradient Echo (MPRAGE) scan for each participant. Image acquisition parameters at each timepoint were as follow: 176 sagittal slices, with a 1mm^3^ voxel size, bandwidth 236 Hz/pixel, FOV=256×256 mm, matrix 256×256, 2300 ms repetition time, and 2.98 echo time and a 9 -degree flip angle.

Prior to pre-processing, all raw T1w images were visually examined for artefacts and then subjected to an automated quality control procedure^49^. Four patient MRI scans did not pass image quality control and were excluded due to excessive head movement (n=1) or image artefacts (n=3). The remaining scans were pre-processed using the longitudinal pipeline of the Computational Anatomy Toolbox^25^ (version r1113) for the Statistical Parametric Mapping 12 (SPM12) software^50^ running in Matlab version 2015a. Briefly, for each participant, the T1w images from all available timepoints were rigidly realigned to correct for differences in head position within-subject, and a subject-specific mean image was calculated and used as a reference in a subsequent realignment of all T1w images across all timepoints. The mean image was segmented into grey matter, white matter, and cerebrospinal fluid, and normalised using the Diffeomorphic Anatomical Registration using Exponentiated Lie algebra algorithm (DARTEL)^50^. The resulting spatial normalisation parameters were then applied to the segmentation of the bias-corrected individual images for all available timepoints, and the resulting native space segmentation was used to calculate overall total intracranial volume and total GMV volume (ml) using FSLstats. The grey matter images were then again realigned to a DARTEL normalised template. Finally, the voxel level intensities were modulated by both the linear and non-linear Jacobian determinants derived from the previous spatial normalisation to preserve the total amount of grey matter. Finally, the resulting modulated and normalised grey matter images were spatially smoothed using an 8mm Gaussian kernel.

### Statistical Analysis – Additional Details

Our secondary analysis included the 12-month follow-up timepoint, with the contrast of interest was a linear polynomial contrast examining differences in linear trend between the three groups. We constrain our contrasts in this way, because our hypotheses concern linear interactions between group and time over the follow-up period (as per Figure 1). The treatment period for the trial ended after 6-months. Thus, in principle, clinicians and patients were no longer bound by the treatment protocol after this point. In practice, four PIPT patients commenced antipsychotic medication in this intervening period, in addition to the four patients who had commenced at the 3-month timepoint. Thus, between the 3-month and 12-month scan, a total of eight PIPT patients commenced antipsychotics, whereas all MIPT patients continued medication with varying degrees of exposure. We thus specified a covariate quantifying cumulative exposure to antipsychotics (olanzapine equivalent, milligrams) for all eight patients within the PIPT group who were exposed to medication at the 3-month or 12-month timepoint. This procedure allowed us to statistically adjust for antipsychotic exposure in the PIPT group when attempting to disentangle the long-term effects of illness and medication exposure on GMV. To ensure that this approach did not substantially influence the results, we repeated the analysis after removing the eight antipsychotic-exposed patients from the PIPT group. The results were largely consistent (see Supplementary Figure 1).

We investigated the potentially confounding effects of DUP, concomitant medication, and substance use on GMV change in each region showing a significant group x time interaction. The effect of DUP prior to recruitment was examined using (1) a one-way ANOVA to examine baseline differences between three DUP strata (0-30 days, 31-90 days, and >90 days); and (2) a two-way ANOVA to assess whether there was a significant interaction effect between time, DUP stratum, and treatment group. To study the effects of non-antipsychotic medications (see Supplementary Table 2), we conducted a three separate two-way ANOVA testing for an interaction between time and percentage patients who received each of the three classes of concomitant medication (benzodiazepines, antidepressants and other psychotropic medication) during the treatment period (the “other” category included people taking zopiclone, dexamethasone, benztropine and clonidine). To assess the effect of substance use, we ran Spearman correlations between regional GMV change and either the WHO-ASSIST total substance use score or the cannabis use sub-score.

We assessed the functional impact of any regions showing a statistically significant group by time interaction by correlating percent change in GMV over time (measured by the first eigenvariate of the region, adjusted for covariates) with percent change in scores on the pre-registered^51^ primary outcomes of the clinical trial; namely the SOFAS and BPRS-4 total scores. Associations were quantified using Spearman’s rank correlations with the threshold for significance set at p<.025 (Bonferroni-adjusted for two comparisons). Additional exploratory correlations between all available clinical and functional scales were Bonferroni-corrected for ten comparisons (p<.005).

#### Results of the Baseline Analysis at an Uncorrected Threshold

At an uncorrected threshold (k > 10, p < 0.001; Supplementary Table 3), patients showed reduced GMV within the right postcentral gyrus, right supramarginal gyrus, right frontal pole, right insula, middle temporal gyrus, and left hippocampus. As expected, no significant total (F = 0.530, p = 0.470) or voxel-level GMV differences were found between the MIPT and PIPT groups at baseline at either corrected or uncorrected thresholds.

#### Results of the Confounding Variables Analyses

The effect of DUP on baseline pallidal volume was not significant (F = .011, p = .918), nor was the interaction between DUP and treatment group on percentage change in pallidal volume between baseline and 3 months (F = .240; p = .628). Similarly, the interactions between treatment group and use of benzodiazepines (F = 1.01; p = .359), antidepressants (F =.552; p = .463) or other psychotropic medication (F = 2.05; p = .163) on percentage change in pallidal volume between baseline and three months were not significant. We also found no significant correlation between percentage change in pallidal volume and total substance use (p = .049; p =.786) or cannabis use (ρ = -.186; ρ = .300).

**Supplementary Table 1.** – Cumulative antipsychotic exposure (in Olanzapine milligram equivalates)

|  | Baseline | 3-months | 12-months |
| --- | --- | --- | --- |
| PIPT, mg (M, SD) | 0.16 (0.59) | 78.5 (216)<sup>1</sup> | 608 (1111) |
| MIPT, mg (M, SD) | 1.03 (5.57) | 420 (248) | 1311 (1011) |

<sup>1</sup> Note: In our primary analysis, the 4 patients within the placebo (PIPT) group who were exposed to antipsychotic medication at amounts greater than the study inclusion criteria at the 3-month timepoint were excluded from the analysis, thus cumulative antipsychotic exposure of the analysis sample was 4.57mg (14.5)

**Supplementary Table 2.** – Percentage of each treatment group who received each class of concomitant medication between baseline and 3-months

|  | Benzodiazepine | Antidepressant | Other<sup>1</sup> |
| --- | --- | --- | --- |
| PIPT, % | 30.0 | 56.7 | 30.0 |
| MIPT, % | 62.0 | 51.7 | 41.4 |

<sup>1</sup> This category included people taking zopiclone, dexamethasone, benztropine and clonidine.

**Supplementary Table 3.** – Regions showing reduced grey matter volume in patients at baseline at an uncorrected threshold (p < .001)

| *Baseline Comparison* | Hemisphere | Peak MNI<sub>spm</sub> coordinates (x,y,z) | Cluster size voxels (mm<sup>3</sup>) |
| --- | --- | --- | --- |
| Supramarginal gyrus | Right | 52.5, -36, 37.5 | 191 (645) |
| Hippocampus | Left | -31.5, -28.5, -12 | 94 (317) |
| Middle-Temporal cortex | Right | 49.5, -58.5, 3 | 93 (314) |
| Frontal pole | Right | 13.5, 60, 7.5 | 57 (192) |
| Postcentral gyrus | Right | 18, -36, 48 | 30 (101) |
| Insula Cortex | Right | 31.5, 15, -1.5 | 21 (71) |

**Supplementary Figure 1.**
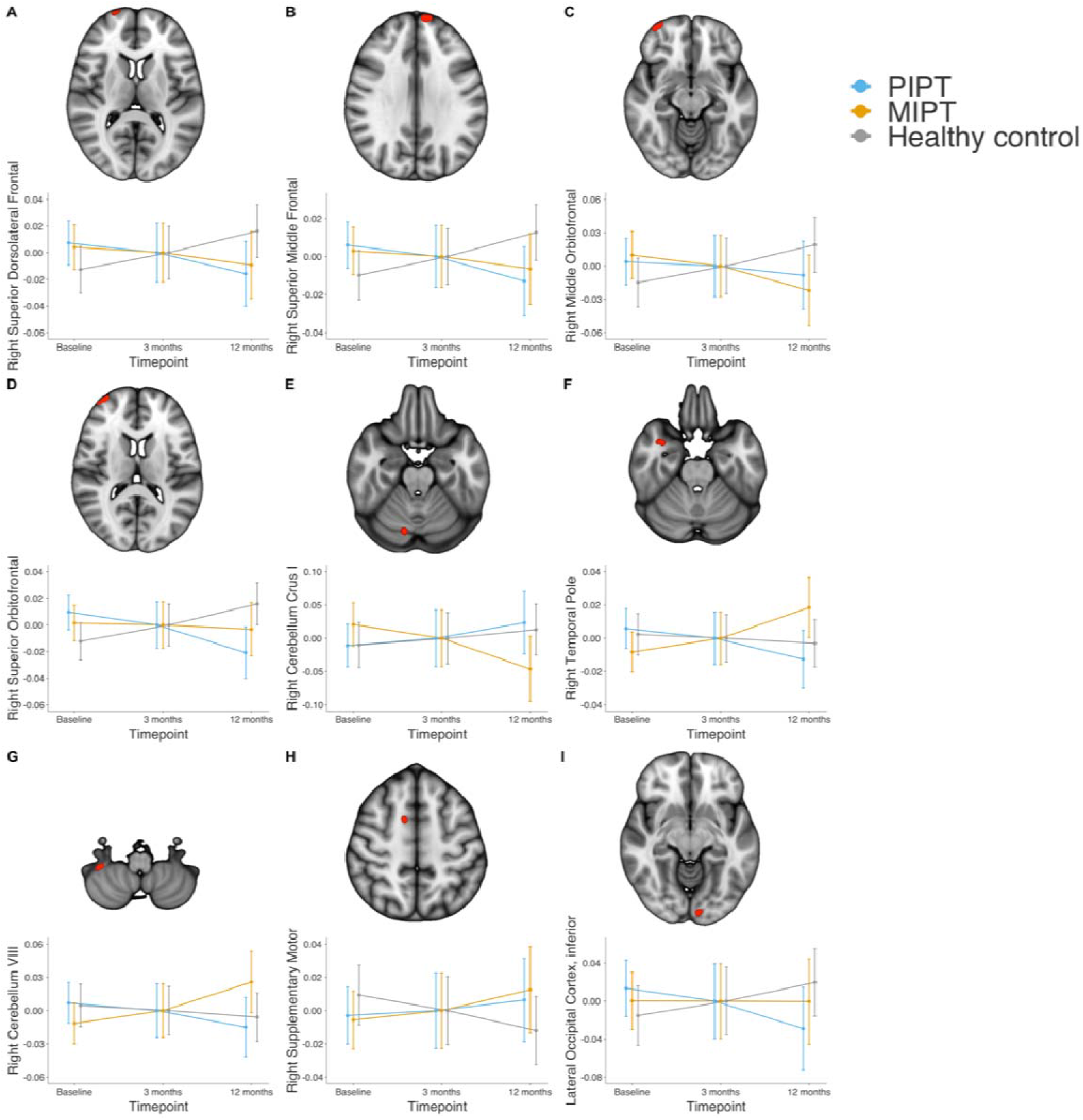
– Red clusters indicate anatomical locations where differences in linear trend (p < 0.001, uncorrected) were detected between baseline and 12 months. In this analysis, patients in the placebo group who were exposed to antipsychotic medication during the 12 months were removed from the analysis. Bottom row of each panel shows the nature of the interaction. Abbreviations: PIPT = placebo plus intensive psychosocial therapy, MIPT = antipsychotic medication plus intensive psychosocial therapy.

**Supplementary Figure 2.**
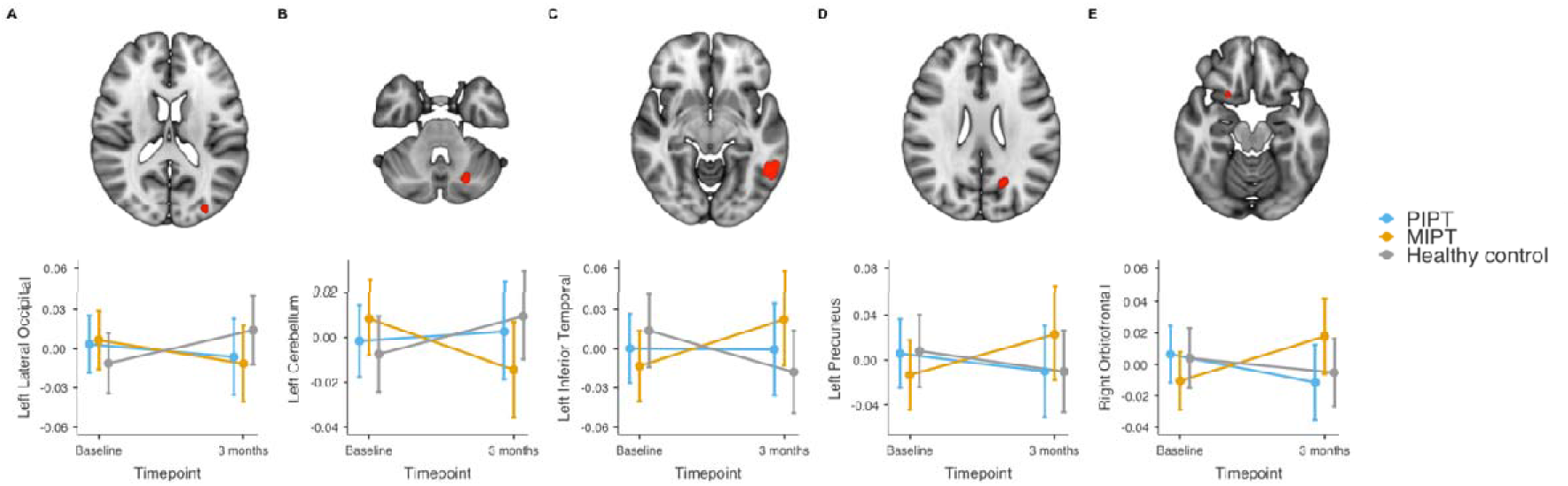
– Red clusters indicate anatomical locations where significant group by time interactions (p < 0.001, uncorrected) were detected between baseline and 3 months. Bottom row shows the nature of the interaction. A) left lateral occipital cortex; B) left cerebellum; C) left inferior temporal cortex; D) left precuneus; E) right orbitofrontal. Abbreviations: PIPT = placebo plus intensive psychosocial therapy, MIPT = antipsychotic medication plus intensive psychosocial therapy.

**Supplementary Figure 3.**
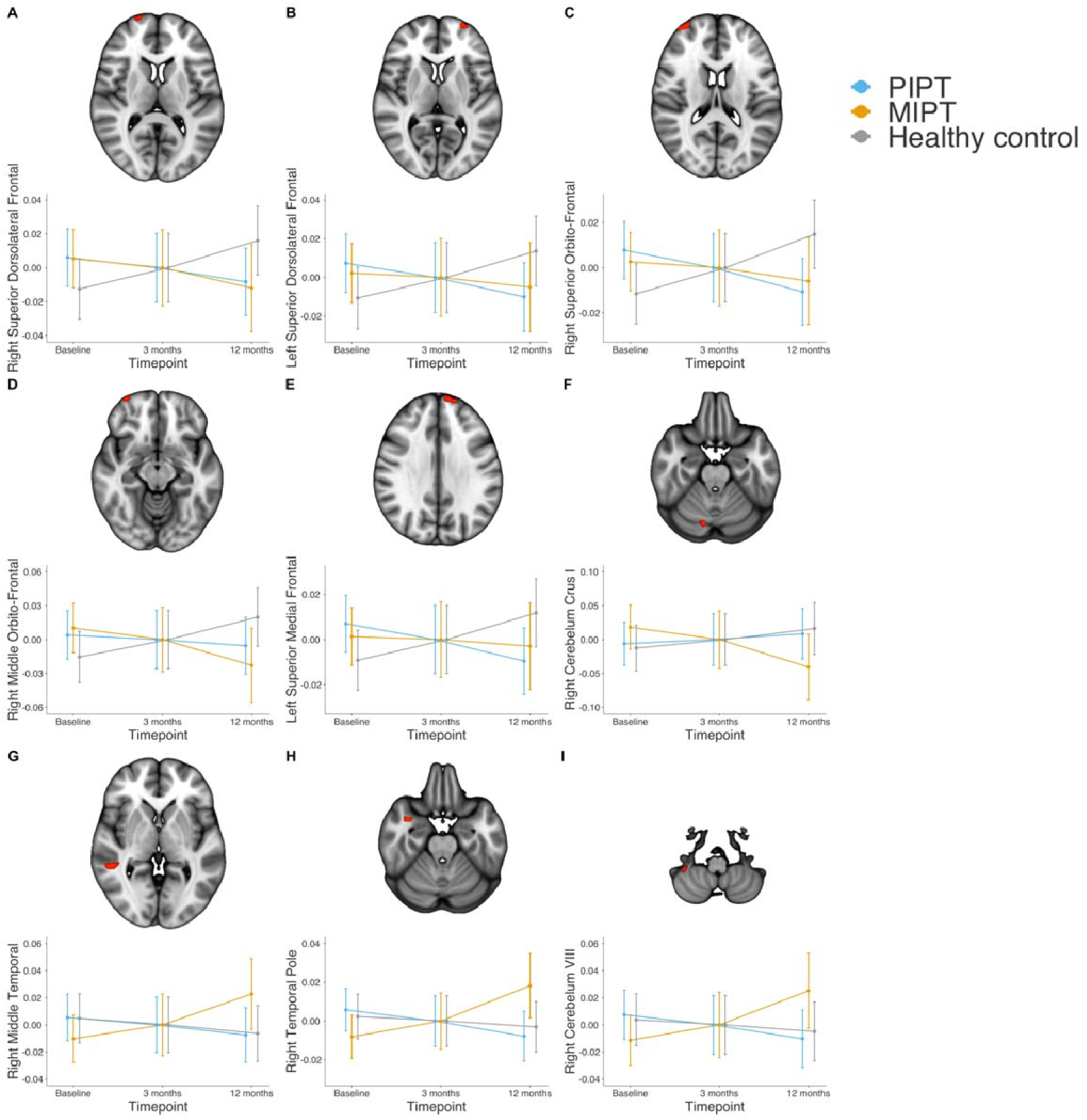
– Red clusters indicate anatomical locations where differences in linear trend (p < 0.001, uncorrected) were detected between baseline and 12 months. Bottom row of each panel shows the nature of the interaction. A) right dorsolateral superior frontal gyrus; B) left dorsolateral superior frontal gyrus; C) right superior orbito-frontal gyrus; D) right middle orbito-frontal gyrus; E) left superior medial frontal gyrus; F) right cerebellar crus I; G) right middle temporal gyrus; H) right temporal pole; I) right cerebellar VIII; Abbreviations: PIPT = placebo plus intensive psychosocial therapy, MIPT = antipsychotic medication plus intensive psychosocial therapy.

**Supplementary Figure 4.**
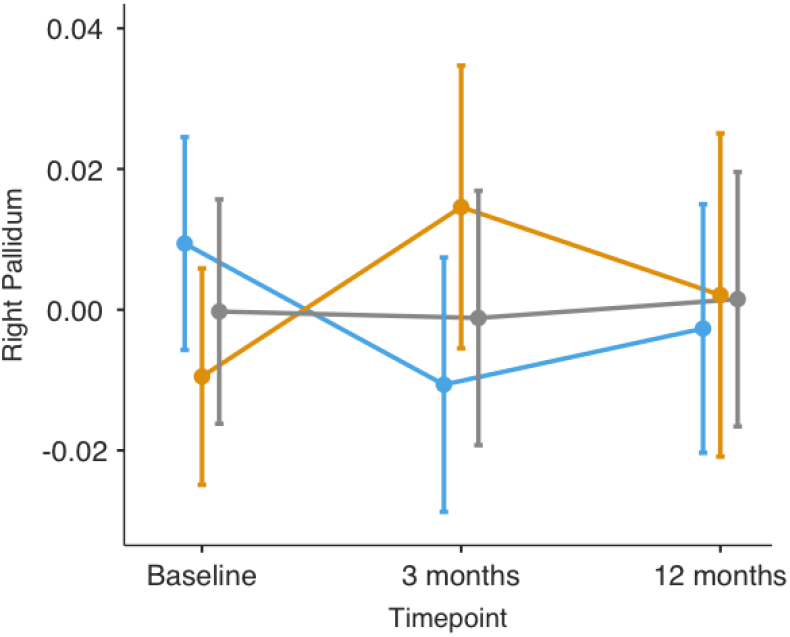
– Volume of pallidal cluster at 12-months follow-up. A) The principal pallidal GMV eigenvariate for each group at baseline and 3-month follow-up, and 12-month follow-up, adjusted for model covariates. The pattern of results remained largely the same after patients in the patients in the PIPT group who were exposed to antipsychotic medication were removed from the analysis.

#### Results from White Matter Analyse at an Uncorrected Threshold

At an uncorrected threshold (k > 10, p < 0.001), interaction effects were detected within the white matter of the right cerebellar lobule V and crus II. These effects were consistent with a putative neurotoxic effect and an unmodified illness-related change, respectively (Supplementary Figure 3b-c).

No significant interactions were detected when including the 12-month time point. At an uncorrected threshold (k > 10, p < 0.001), an interaction effect consistent with a unmodified illness-related change was detected within left frontal white matter (Supplementary Figure 4).

**Supplementary Figure 5.**
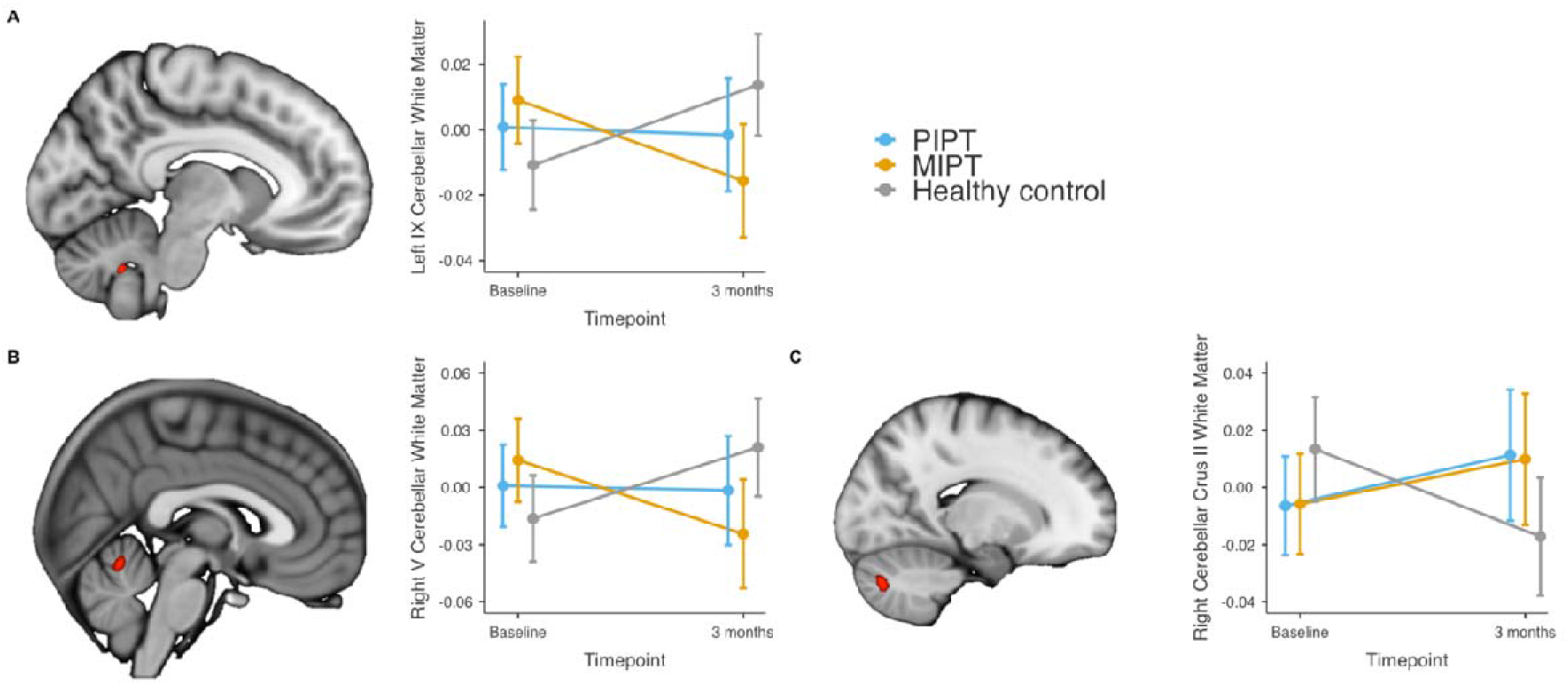
– The principal cerebellar white matter volume eigenvariate for each group at baseline and 3-month follow-up, adjusted for model covariates. Error bars show 95% confidence intervals. A) Cerebellar lobule IX cluster (p < 0.05, FWE-corrected) B) cerebellar lobule V (p < 0.001, uncorrected), C) Cerebellar crus II (p < 0.001, uncorrected)

**Supplementary Figure 6.**
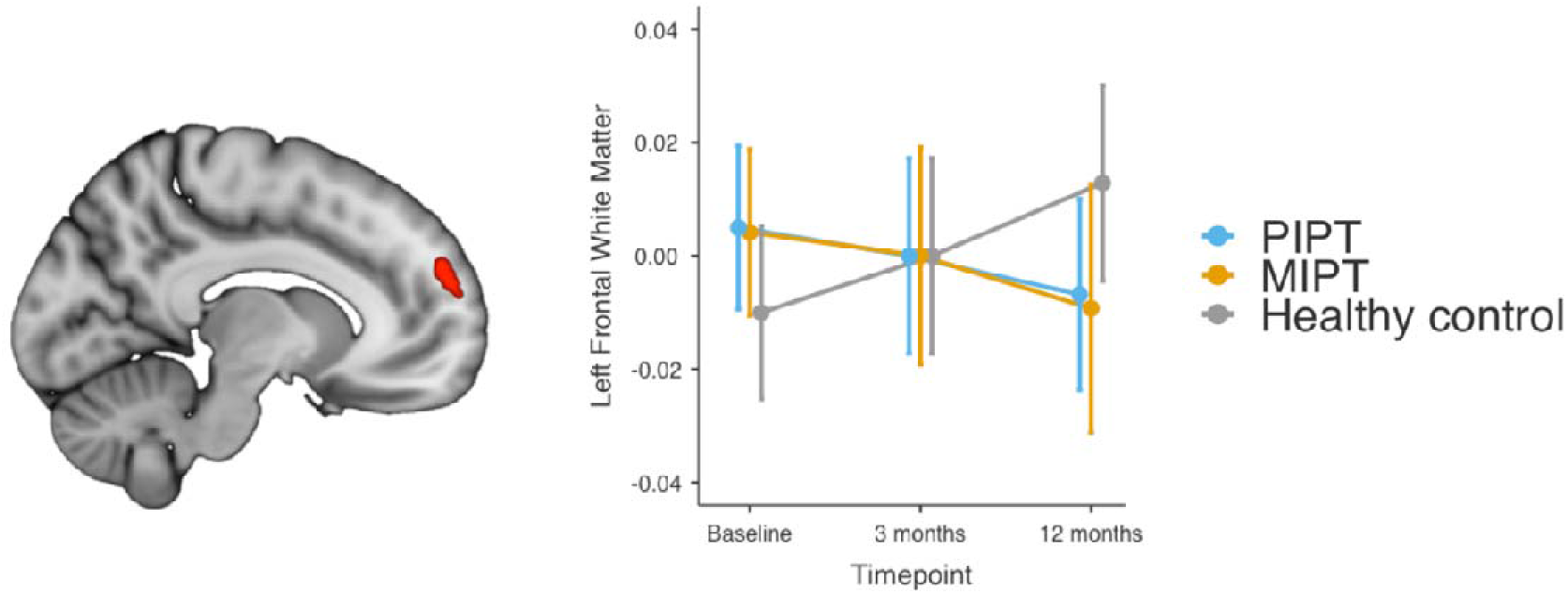
– Red clusters indicates frontal white matter where differences in linear trend (p < 0.001, uncorrected) were detected between baseline and 12 months.

## Notes

### Competing Interest Statement

Conflict of interest disclosures
None reported, to be finalised

### Clinical Trial

ACTRN12607000608460

### Funding Statement

Funding and medication support that was provided to this study by Janssen-Cilag, Australia (2007-2012) as an unrestricted investigator-initiated grant. The study was supported by grant 1064704 from the Australian National Health and Medical Research Council.
Role of the Funder/Sponsor
The funding source had no role in the design or conduct of the study: collection, management, analysis, and interpretation of data; preparation, review or approval of the manuscript; and decision to submit the manuscript for publication.

